# Alcohol Use Disorder Is Associated with Higher Risks of Adverse Brain Outcomes

**DOI:** 10.1101/2022.05.04.22274661

**Authors:** Pengyue Zhang, Howard Edenberg, John Nurnberger, Dongbing Lai, Feixiong Cheng, Yunlong Liu

## Abstract

Alcohol use disorder (AUD) is on the ascendancy in the US older adult population, while the association between AUD and adverse brain outcomes remains inconclusive. The objective of this work is to investigate the associations between AUD with the onset of Alzheimer’s disease and Parkinson’s disease. In a retrospective cohort design using US insurance claim data (2007-2020), 129,182 patients with AUD were matched with 129,182 controls by age, sex, race, and clinical characteristics. After adjusting for covariates, AUD was associated with a higher risk of Alzheimer’s disease (female adjusted HR=1.78, 95% CI: 1.68-1.90, P<0.001; male adjusted HR=1.80, 95% CI: 1.71-1.91, P<0.001) and a higher risk of Parkinson’s disease (female adjusted HR=1.49, 95% CI: 1.32-1.68 P<0.001; male adjusted HR=1.42, 95% CI: 1.32-1.52, P<0.001) in the overall sample. In separate analyses of Black, White, and Hispanic patients, those with AUD had higher risk of Alzheimer’s disease (adjusted HRs≥1.58; Ps≤0.001). A significantly elevated risk for Parkinson’s disease was found only in the White subpopulation (female adjusted HR=1.55, 95% CI: 1.36-1.77, P<0.001; male adjusted HR=1.45, 95% CI: 1.33-1.57, P<0.001). Alcohol use disorder is associated with Alzheimer’s disease. Alcohol use disorder is associated with Parkinson’s disease in White persons. Cognitive screening and neurological examination among older adults with severe problematic alcohol use hold the promise for early detection of Alzheimer’s disease and Parkinson’s disease.

## 1. Background

Alcohol drinking is common among the US older adult population aged ≥65 years. US nation-wide survey data from 2017 - 2019 has shown that: 1) over 40% of US older adults used alcohol; and 2) over 13% of US older adults engaged in binge or heavy alcohol use [1, 2]. The prevalence of alcohol use disorder (AUD) in the older adult population (age ≥65 years) increased from 3.2% (95% confidence interval [CI]: 2.6%-4.0%) to 5.6% (95% CI: 4.8%-6.6%) from years 2001-2002 to 2012-2013, a 1.75-fold increase (P<0.05) [3].

Despite the fact that excessive alcohol consumption is known to have neurotoxic consequences [4, 5], population-based association studies investigating adverse brain outcomes for alcohol users and/or patients with AUD have shown mixed results [6]. AUD was associated with increased risk of dementia including Alzheimer’s disease (hazard ratio [HR]=3.34, P<0.0001) in an analysis of 31.6 million French patients [7]. Based on the aforementioned study and literature reviews, excessive alcohol consumption (>21 units/week) was considered as a new risk factor for dementia in the 2020 report of the Lancet Commission on Dementia Prevention, Intervention, and Care [8]. AUD was also associated with an increased risk of Parkinson’s disease (HR=1.38, P<0.0001) among 602,930 Swedish patients with an average of 13 years of follow-up [9]. Additionally, an over 30-year observation study in England concluded that both moderate and high alcohol consumption (i.e. 14-21 and >21 units/week) increased the risk (odds ratios [ORs]>3.4, Ps<0.007) for developing hippocampal atrophy [10]. On the other hand, some studies found that alcohol use and AUD had no association or were associated with decreased risk of degenerative brain diseases. For instance, a 3-year observational study in Germany found that alcohol users had reduced risk of overall dementia (HR=0.71, P=0.028) and Alzheimer disease (HR=0.58, P=0.013) [11]. A cohort study of middle-aged and older US adults (N=19,887) concluded a U-shaped relationship between weekly alcohol consumption and cognitive function with better cognitive function at 10-14 drinks per week [12]. Another 13-year longitudinal study of a large US cohort (N=132,403) identified no association between alcohol intake and risk of Parkinson disease [13].

Currently, AUD is on the ascendancy in the older adult population [3]. Thus, more real-world evidence on adverse brain outcomes for patients with AUD is warranted [14]. In this study, we investigated the association between AUD and Alzheimer dementia and Parkinson disease using a large-scale US insurance claim dataset.

## 2. Methods

### 2.1 Data

We used the OPTUM Clinformatics Data (2007Q1-2020Q4). The data are derived from commercially insured US individuals and US Medicare Advantage beneficiaries. The data included individual level demographics (e.g., sex, birth year, and race), monthly enrollment records, medical claims (e.g., emergency department [ED] visits, inpatient hospitalizations, and outpatient visits), procedure claims, pharmacy claims, laboratory results, and death records.

### 2.2 Data Accessibility and Institutional Review Board (IRB)

Data accessibility is granted by the license agreement between Indiana University and OPTUM. The OPTUM Clinformatics Data is de-identified. The Indiana University Institutional Review Board (IRB) designated this study as exempt.

### 2.3 Phenotyping algorithm

We used ICD-9 and ICD-10 codes to identify phenotypes. Specifically, we used alcohol related diagnosis codes defined by the US Centers for Disease Control and Prevention (CDC) to identify alcohol use disorders (ICD-9: 291*, 303*, 305.0*, 357.5, 425.5, 535.3, 571.0, 571.1, 571.2, 571.3; ICD-10: F10*, G62.1, G31.2, G72.1, I42.6, K29.2, K70*, K85.2, K86.0, Q86.0) [15]. We used algorithms from the published literature to identify Alzheimer’s disease (ICD-9 331.0; ICD-10: F00*, G30*) [16] and Parkinson’s disease (ICD-9: 332*, 331.82, 333.0; ICD-10: G20, G21*, G31.83, G31.85, G23.1, G23.2, G90.3) [17]. We used the R package *comorbidity* [18] to identify dementia, heart disease (myocardial infarction and congestive heart failure), cerebrovascular disease, chronic pulmonary disease, liver disease, diabetes, renal disease, HIV infection, hemiplegia or paraplegia, and malignancy. We used the algorithm derived by Chen *et al*. [19] to identify fall, and hypertension. We used the algorithm derived by Quan *et al*. [20] to identify depression. We used the algorithm defined in Anna Nordström and Peter Nordström to identify traumatic brain injury [21].

### 2.4 Study design

We used a cohort design to compare the AUD patient group and a matched control group with respect to adverse brain outcomes. The study design was illustrated in Figure 1. We included individuals that were: 1) ≥60 years on the first day of enrollment; and 2) continuously enrolled for ≥3 years. We identified 251,321 patients with AUD. Among these, we excluded 111,528 patients with AUD that were: 1) diagnosed with AUD within 365 days of enrollment; and 2) diagnosed with Alzheimer’s disease, cancer, dementia, hemiplegia/paraplegia, HIV infection, and/or Parkinson’s disease prior to the diagnosis of AUD. After exclusion, 139,793 were eligible to be matched. We matched patients with AUD and controls on age, sex, race, and comorbidities common among older adults and/or associated with AUD and adverse brain outcomes [8, 22]. Specifically, we matched on the following conditions: cerebrovascular disease, chronic pulmonary disease, depression, diabetes, fall, heart disease (myocardial infarction and congestive heart failure), hypertension, liver disease, renal disease, and traumatic brain injury. For AUD patients, the index date was the diagnosis date for AUD. For each matched pair, the control index date was chosen to be the corresponding AUD patient’s index date (Figure S1). To summarize, individuals within a matched pair: 1) were not diagnosed with Alzheimer’s disease, cancer, dementia, hemiplegia/paraplegia, HIV infection, and/or Parkinson’s disease prior to the index date; and 2) had the same demographics (age, race and sex) and comorbidity status on the index date. The final data set included 129,182 patients with AUD and 129,182 matched control individuals.

**Figure 1.**
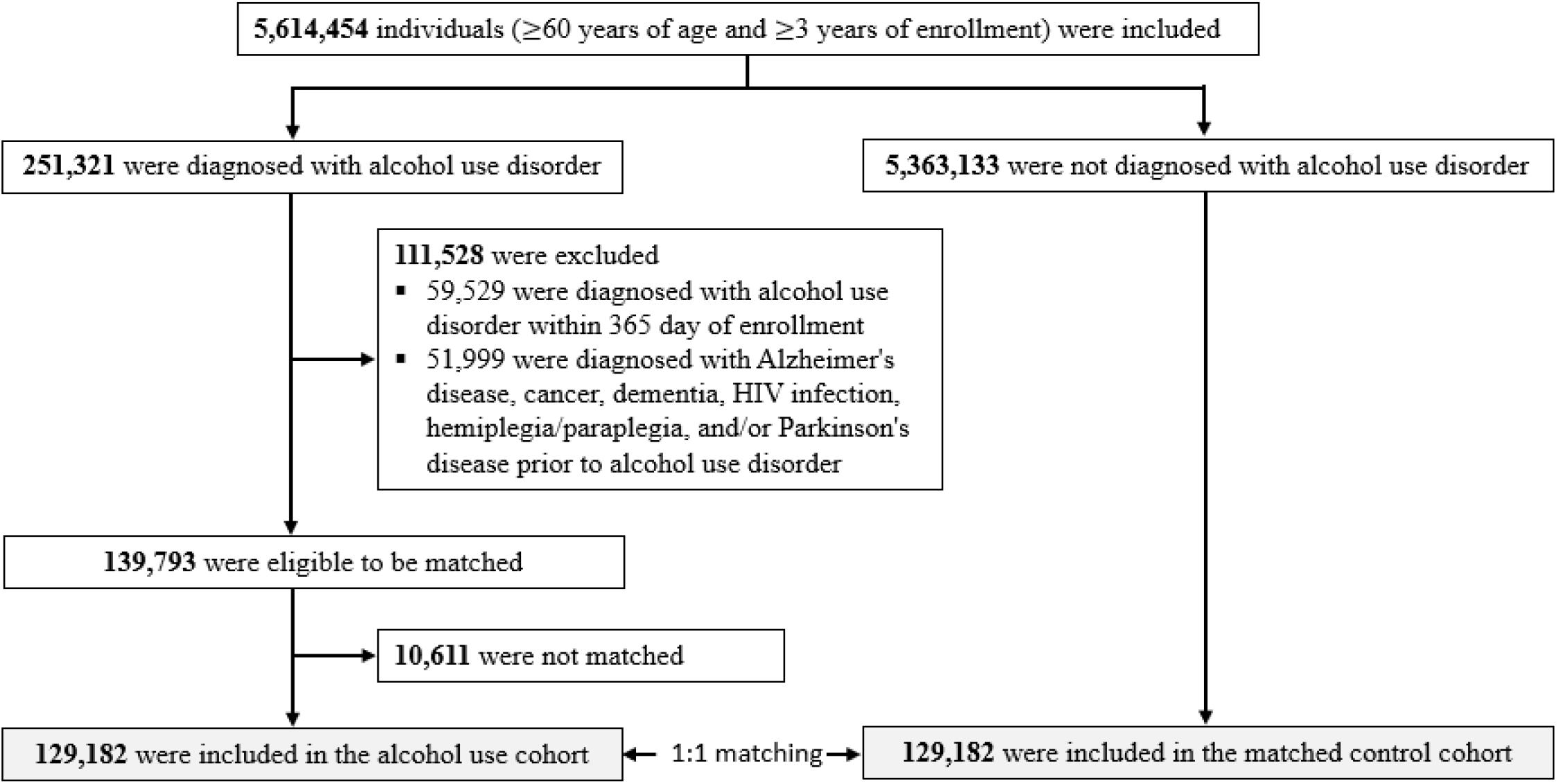
Study population and cohort matching process.

### 2.5 Dependent variable

The primary outcomes were time from index date (i.e., day-0) to the earliest diagnosis date of Alzheimer’s disease or Parkinson’s disease (Figure S1). The secondary outcome was time from index date to the earliest diagnosis of dementia. For all outcomes, the earliest diagnosis date is defined as the first diagnosis date after the index date. Individuals without any outcome were censored at the last month of enrollment or on 12/31/2020, whichever came first.

### 2.6 Statistical analysis

We used a Cox proportional hazard model to estimate covariate-adjusted hazard ratios (HRs), 95% confidence intervals (CIs), and p-values [23]. We used the Kaplan–Meier estimator to estimate cumulative incidence curves for the patients with AUD and the matched controls [24]. We used the log-rank test to compare the incidence rates over time between the patients with AUD and the matched controls [25]. The Cox proportional hazards models were adjusted for the following covariates: age (60-74 and 75-100), race (White, Hispanic, Black, Asian, and unknown), cerebrovascular disease (yes/no), chronic pulmonary disease (yes/no), depression (yes/no), diabetes (yes/no), fall (yes/no), heart disease (yes/no), liver disease (yes/no), renal disease (yes/no), and traumatic brain injury (yes/no). All analyses were stratified by sex because men and women have different incidence of AUD and adverse brain outcomes [26, 27].

Additionally, besides analyzing the whole dataset (N=258,364), we also did sensitivity analyses in: 1) race-specific subgroups, and 2) patients with alcohol abuse (i.e. ICD-9/10: 305.0* and F10.1*) and the matched controls. In all sensitivity analyses, we conducted Cox proportional hazard regression, computed Kaplan–Meier estimator, and conducted log-rank test. In secondary analysis, we investigated the association between AUD and dementia, as well as the cumulative incidence of dementia for patients with AUD and the matched controls after index date. All analyses were performed with R 4.0.2.

## 3. Results

The whole study dataset included 258,364 individuals (129,182 patients with AUD and 129,182 matched control individuals). Table 1 shows the demographic characteristics of the AUD patient group and the matched control group at baseline (i.e. the index dates). Demographics of subpopulations are presented in Supplementary Material Table S1 and S4.

**Table 1.** Demographics of the study population (N=258,364)

|  | Female (N=90,486) |  |  |  | Male (N=167,878) |  |  |  |
| --- | --- | --- | --- | --- | --- | --- | --- | --- |
|  | Alcohol use disorder |  | Matched Control |  | Alcohol use disorder |  | Matched Control |  |
| Age group |  |  |  |  |  |  |  |  |
| 65-75 | 30,511 | 67.40% | 30,511 | 67.40% | 61,190 | 72.90% | 61,190 | 72.90% |
| 76-100 | 14,732 | 32.60% | 14,732 | 32.60% | 22,749 | 27.10% | 22,749 | 27.10% |
| Race |  |  |  |  |  |  |  |  |
| White | 34,230 | 75.70% | 34,230 | 75.70% | 59,904 | 71.40% | 59,904 | 71.40% |
| Hispanic | 3,605 | 8% | 3,605 | 8% | 10,221 | 12.20% | 10,221 | 12.20% |
| Black | 4,147 | 9.20% | 4,147 | 9.20% | 7,617 | 9.10% | 7,617 | 9.10% |
| Asian | 676 | 1.50% | 676 | 1.50% | 1,652 | 2% | 1,652 | 2% |
| Unknown | 2,585 | 5.70% | 2,585 | 5.70% | 4,545 | 5.40% | 4,545 | 5.40% |
| Comorbidity |  |  |  |  |  |  |  |  |
| Cerebrovascular disease | 9,315 | 20.60% | 9,315 | 20.60% | 16,294 | 19.40% | 16,294 | 19.40% |
| Depression | 16,655 | 36.80% | 16,655 | 36.80% | 14,792 | 17.60% | 14,792 | 17.60% |
| Diabetes | 11,697 | 25.90% | 11,697 | 25.90% | 27,258 | 32.50% | 27,258 | 32.50% |
| Fall | 8,552 | 18.90% | 8,552 | 18.90% | 8,069 | 9.60% | 8,069 | 9.60% |
| Heart disease | 7,984 | 17.60% | 7,984 | 17.60% | 18,848 | 22.50% | 18,848 | 22.50% |
| Hypertension | 35,028 | 77.40% | 35,028 | 77.40% | 65,923 | 78.50% | 65,923 | 78.50% |
| Liver disease | 7,350 | 16.20% | 7,350 | 16.20% | 11,258 | 13.40% | 11,258 | 13.40% |
| Chronic pulmonary disease | 18,136 | 40.10% | 18,136 | 40.10% | 29,046 | 34.60% | 29,046 | 34.60% |
| Renal disease | 7,625 | 16.90% | 7,625 | 16.90% | 14,770 | 17.60% | 14,770 | 17.60% |
| Traumatic brain injury | 695 | 1.50% | 695 | 1.50% | 629 | 0.70% | 629 | 0.70% |

Figure 2 shows the covariate adjusted hazard ratios (HRs) for patients with AUD relative to matched controls. For Alzheimer’s disease, both female and male patients with AUD were associated with higher risk in the whole study population (female adjusted HR=1.78 [95% CI: 1.68-1.90]; male adjusted HR=1.80 [95% CI: 1.71-1.91]; Ps<1 × 10^−8^). In race-specific subgroup analyses, female patients with AUD were associated with higher risk of Alzheimer’s disease (adjusted HRs≥1.58; Ps≤2 × 10^−S^) in White/Hispanic/Black subpopulations; male patients with AUD were associated with higher risk of Alzheimer’s disease (adjusted HRs≥1.59; Ps≤0.02) in all race-specific subgroups. White female patients with AUD had comparable risks for Alzheimer’s disease to White male patients with AUD. Hispanic and Black female patients with AUD had higher risks for Alzheimer’s disease than corresponding male patients with AUD. For Parkinson’s disease, both female and male patients with AUD were associated with higher risk in the whole study population (female adjusted HR=1.49 [95% CI: 1.32-1.68]; male adjusted HR=1.42 [95% CI: 1.32-1.52]; Ps<1 × 10^−8^). In subgroup analysis, only White patients with AUD were associated with higher risk of Parkinson’s disease (female adjusted HR=1.55 [95% CI: 1.36-1.77]; male adjusted HR=1.45 [95% CI: 1.33-1.57]; Ps< 1 × 10^−8^). White female patients with AUD had higher risk for Parkinson’s disease relative to White male patients with AUD. Results from multivariate Cox analysis of Alzheimer’s disease and Parkinson’s disease are given in Supplementary Material Table S2 and S3.

**Figure 2.**
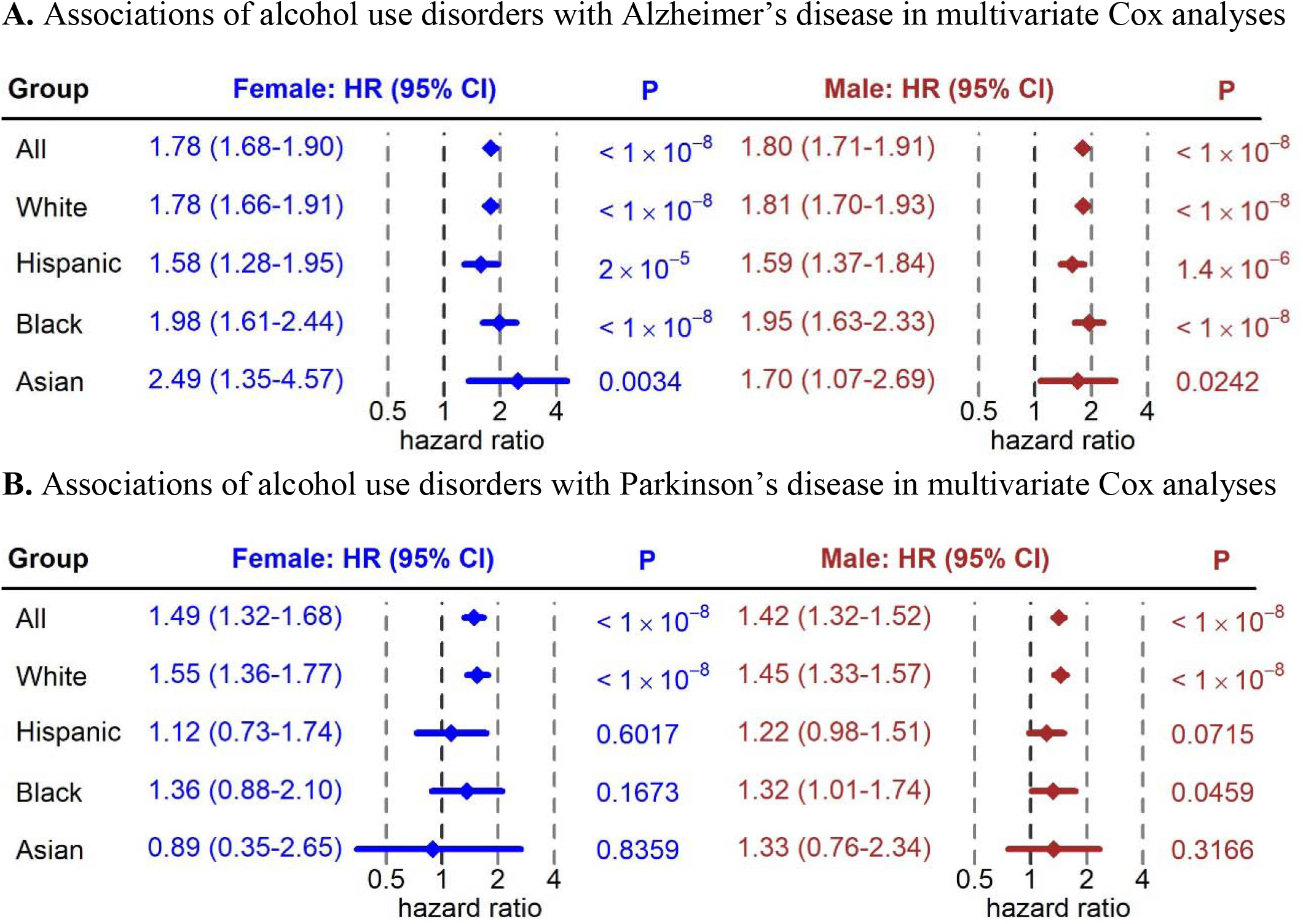
**A**. Associations of alcohol use disorders with Alzheimer’s disease in multivariate Cox analyses (N=258,364). **B**. Associations of alcohol use disorders with Parkinson’s disease in multivariate Cox analyses (N=258,364).

Figure 3 presents the sex-specific cumulative incidence of Alzheimer’s disease after diagnosis date or matched control index date. White/Hispanic/Black female patients with AUD had significant higher cumulative incidence of Alzheimer’s disease than matched controls. Black female patients with AUD had the highest 5-year cumulative incidence (8.4%), followed by White female patients with AUD (8.0%), and then Hispanic female patients with AUD (7.4%). Male patients with AUD had a significantly higher cumulative incidence of Alzheimer’s disease than matched controls in all race-specific subgroups. Black male patients with AUD had the highest 5-year cumulative incidence (6.2%), followed by White male patients with AUD (5.4%), then Hispanic male patients with AUD (5.0%), and last Asian male patients with AUD (4.0%).

**Figure 3.**
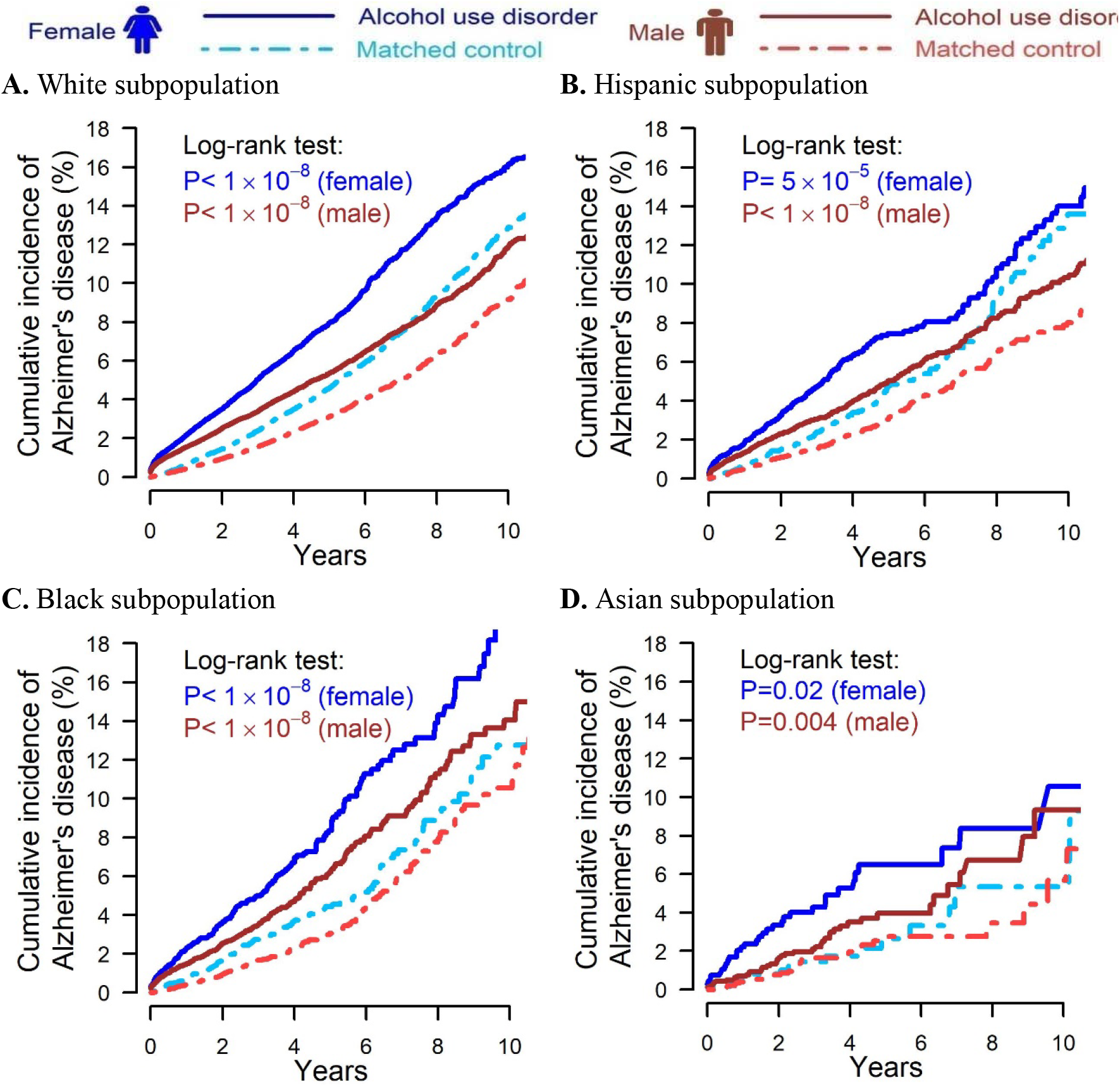
Cumulative incidence of Alzheimer’s disease for patients with alcohol use disorder and matched controls after index date; **A**. White subpopulation (female N=68,460, male N=119,808); **B**. Hispanic subpopulation (female N=7,210, male N=20,442); **C**. Black subpopulation (female N=8,294, male N=15,234); **D**. Asian subpopulation (female N=1,352, male N=3,304).

Figure 4 presents the sex-specific cumulative incidence of Parkinson’s disease after diagnosis date or matched control index date. Both White female and male patients with AUD had a significantly higher cumulative incidence of Parkinson’s disease compared with matched controls. White female patients with AUD had a 2.1% 5-year cumulative incidence, while matched controls had a 1.5% 5-year cumulative incidence. White male patients with AUD had a 3.0% 5-year cumulative incidence, while matched controls had a 2.1% 5-year cumulative incidence.

**Figure 4.**
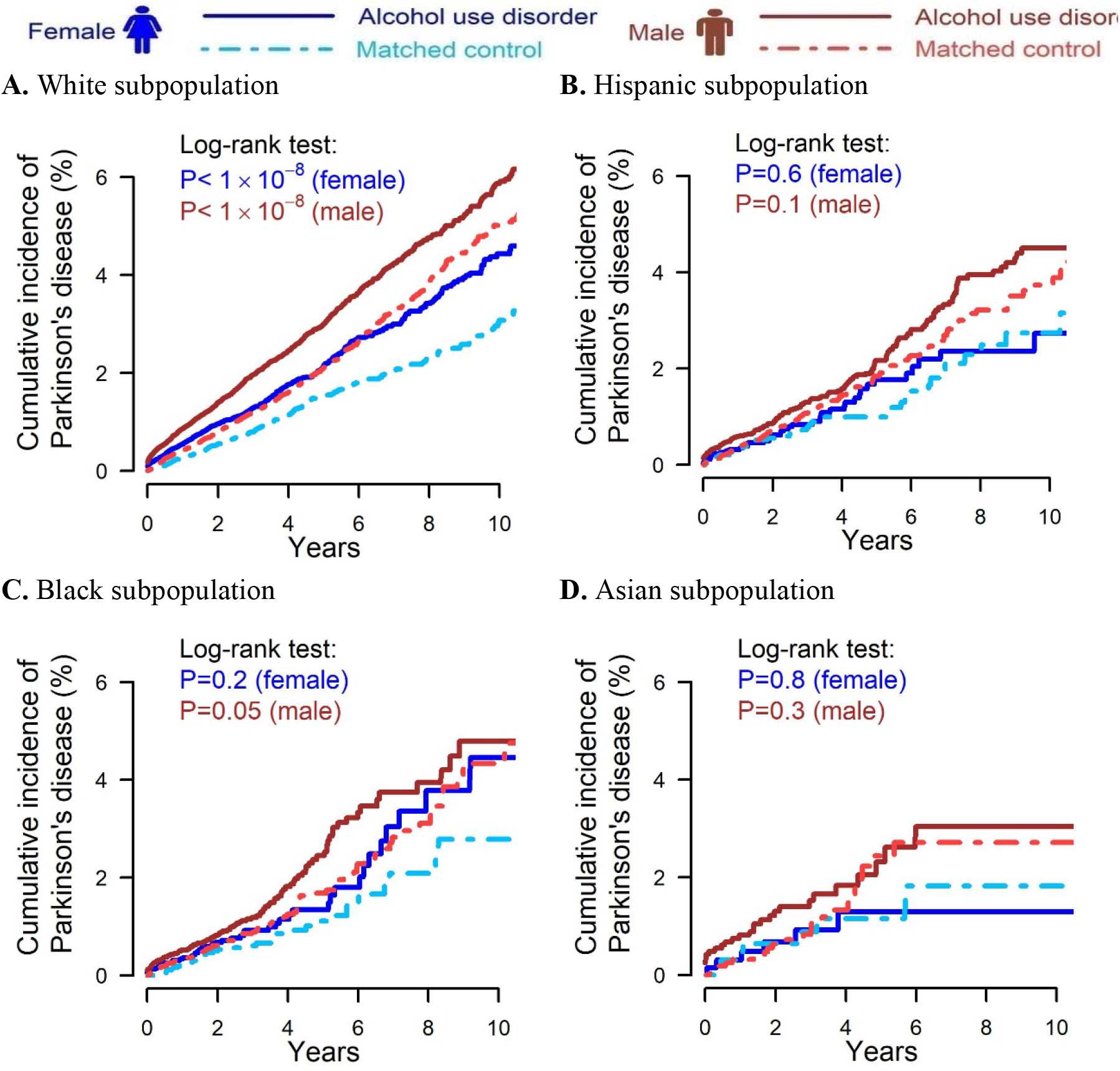
Cumulative incidence of Parkinson’s disease for patients with alcohol use disorder and matched controls after index date; **A**. White subpopulation (female N=68,460, male N=119,808); **B**. Hispanic subpopulation (female N=7,210, male N=20,442); **C**. Black subpopulation (female N=8,294, male N=15,234); **D**. Asian subpopulation (female N=1,352, male N=3,304).

Secondary and sensitivity analyses are presented in the supplementary figures and tables. AUD was associated with higher risks of dementia in the whole study population and all race-specific subpopulations (adjusted HRs≥1.74, Ps<0.001 [Figure S3]). Alcohol abuse was associated with: 1) higher risk of Alzheimer’s disease in the whole study population (female adjusted HR=1.82 [95% CI: 1.70-1.99]; male adjusted HR=1.80 [95% CI: 1.66-1.95]; Ps≤1 × 10^−8^ [Figure S6]); and 2) higher risk of Parkinson’s disease in the whole study population (female adjusted HR=1.49 [95% CI: 1.25-1.77]; male adjusted HR=1.39 [95% CI: 1.25-1.55]; Ps≤1 × 10^−5^[Figure S6])

## 4. Discussion

This study highlights real-world evidence on adverse brain outcomes following diagnosis of alcohol use disorder (AUD). We found that AUD was associated with higher risks of Alzheimer’s disease and dementia consistently in the whole study population and White/Hispanic/Black subpopulations. We also found that AUD was associated with higher risks of Parkinson’s disease in the whole study population and the White subpopulation.

The primary aim of the study was to investigate the associations between AUD and adverse brain outcomes. Our findings are in agreement with most of the literature [7, 9]. Our race-specific subgroup analysis results provide novel race-specific real-world evidence on adverse brain outcomes following diagnosis of AUD. Pre-clinical and clinical studies have identified mechanisms of AUD-induced neurodegeneration [6]. Specifically, alcohol exposure could induce oxidative stress in brain, hyperglutamatergic excitotoxicity, and neuroinflammation, all of which may subsequently trigger neuronal apoptosis. Thus, our findings together with alcohol’s neurotoxicity suggest cognitive screening and neurological examination among older adults with severe problematic alcohol use holds the promise for early detection of adverse brain outcomes [6].

This study has several limitations. First, the analytical data set only includes commercially insured individuals and US Medicare Advantage beneficiaries. In 2020, Medicare Advantage beneficiaries reached 24 million (i.e., 40% of all Medicare beneficiaries), which represents a geographically and racially diverse older adult population. The study results could be generalized to Medicare Advantage beneficiaries. However, the results may not be generalizable to traditional Medicare beneficiaries, as most Medicare Advantage enrollees have access to additional benefits that not covered by traditional Medicare [28].

Second, administrative claims data is not intended for research purposes. Socioeconomic status and education level are not explicitly measured in the analytical data and these may have significant impact on the outcomes. Further, claims data only represents AUD of individuals when such a diagnosis is present, as well as individuals with other phenotypes. Accordingly, we could not account for phenotypes without a diagnosis. In the analytical data set, the prevalence of AUD is 4.3%, which are likely to be underestimated based on epidemiologic studies [3]. Other possible confounders that we were unable to measure include but not limited to neurological exams, cognitive tests, brain imaging studies, and genetic factors.

A third notable limitation, also associated with the constraints of administrative data, is that we could not observe the alcohol consumption records. We can assume that an AUD diagnosis is associated with higher alcohol consumption prior to the diagnosis. In the US, 23% of older adults engaged in risky drinking, which is defined as consuming 15 or more standard drinks per week or 5 or more on an occasion [29]. Additionally, continuously drinking after diagnosis of AUD and/or relapse of drinking after abstinence are common in the US [30]. Thus, the observed associations between AUD and adverse brain outcomes are likely to be induced by risky drinking. Future studies are warranted to illuminate the mechanism of alcohol induced adverse brain outcome.

In summary, we found that AUD was associated with an increased risk of Alzheimer’s disease and Parkinson’s disease in the whole study population. AUD was associated with the highest risk of Alzheimer’s disease in the Black subpopulation, followed by the White subpopulation, and the Hispanic subpopulation. AUD was associated with increased risk of Parkinson’s disease in the White subpopulation. Female patients with AUD had higher or comparable risks of adverse brain outcomes relative to male patients with AUD.

## Data Availability

Claims data are available from OPTUM, Inc.

