## Supplementary Material for "Alcohol Use Disorder Is Associated with Higher Risks of Adverse Brain Outcomes"

**Figure S1** Illustration of study design.

**Figure S2.** Cumulative incidence of dementia for alcohol use disorder patients and matched controls; **A.** All Races (female N=90,486, male N=167,878); **B.** White subpopulation (female N=68,460, male N=119,808); **C.** Hispanic subpopulation (female N=7,210, male N=20,442); **D.** Black subpopulation (female N=8,294, male N=15,234); **E.** Asian subpopulation (female N=1,352, male N=3,304).

**Figure S3** Association of alcohol use disorders with dementia in multivariate Cox analyses (N=258,364).

**Figure S4.** Cumulative incidence of Alzheimer’s disease for patients with alcohol abuse and matched controls; **A.** All Races (female N=42,164, male N=77,884); **B.** White subpopulation (female N=32,418, male N=56,468); **C.** Hispanic subpopulation (female N=2,768, male N=7,574); **D.** Black subpopulation (female N=4,194, male N=8,408); **E.** Asian subpopulation (female N=476, male N=1,364).

**Figure S5.** Cumulative incidence of Parkinson's disease for patients with alcohol abuse and matched controls; **A.** All Races (female N=42,164, male N=77,884); **B.** White subpopulation (female N=32,418, male N=56,468); **C.** Hispanic subpopulation (female N=2,768, male N=7,574); **D.** Black subpopulation (female N=4,194, male N=8,408); **E.** Asian subpopulation (female N=476, male N=1,364).

**Figure S6.** Association of alcohol abuse with Alzheimer’s disease and Parkinson's disease in multivariate Cox analyses (N= 120,048).

**Figure S7.** Cumulative incidence of dementia for patients with alcohol abuse and matched controls; **A.** All Races (female N=42,164, male N=77,884); **B.** White subpopulation (female N=32,418, male N=56,468); **C.** Hispanic subpopulation (female N=2,768, male N=7,574); **D.** Black subpopulation (female N=4,194, male N=8,408); **E.** Asian subpopulation (female N=476, male N=1,364).

**Figure S8.** Association of alcohol abuse with dementia in multivariate Cox analyses (N=120,048).

**Table S1.** Demographics of race-specific subpopulations of patients with alcohol use disorder.

**Table S2.** Results from multivariate Cox analysis: association with Alzheimer’s disease (N=258,364).

**Table S3.** Results from multivariate Cox analysis: association with Parkinson's disease (N=258,364).

**Table S4.** Demographics of alcohol abuse patients and matched controls.

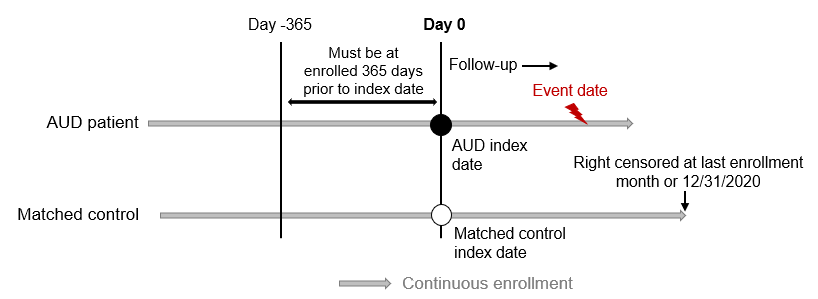

**Figure S1** Illustration of study design.

| 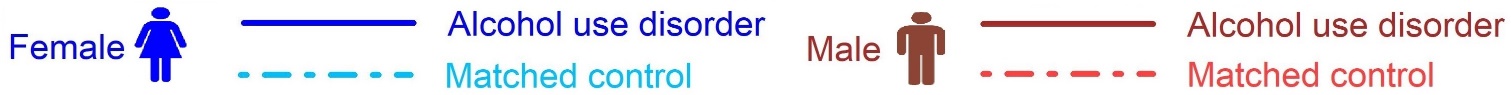 | |
| --- | --- |
| **A.** All Races | |
| 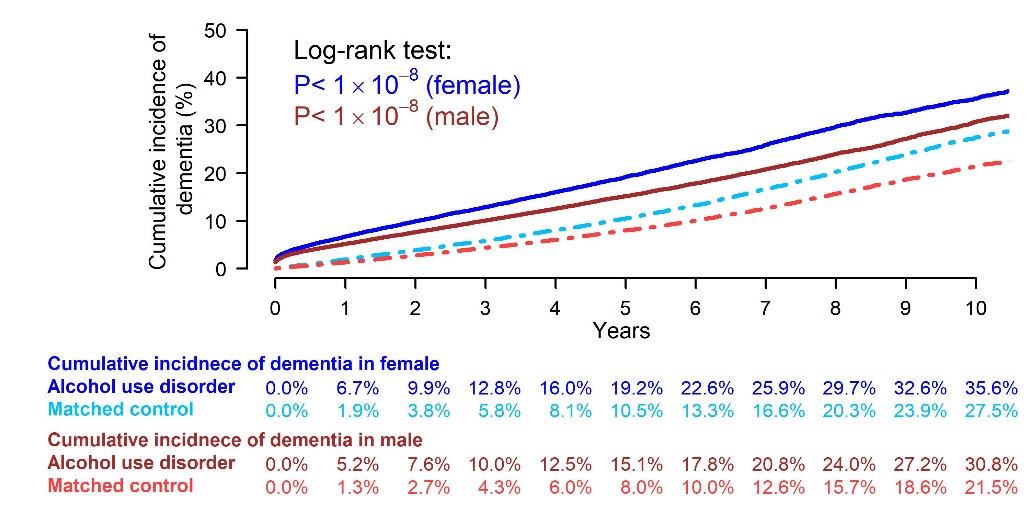 | |
| **B.** White subpopulation | **C.** Hispanic subpopulation |
| 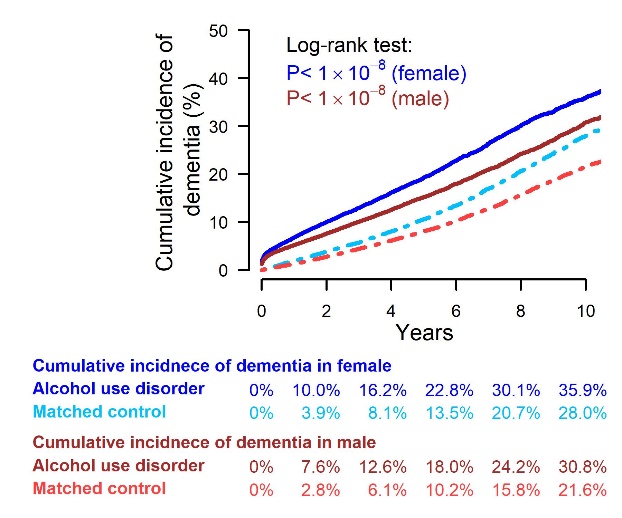 | 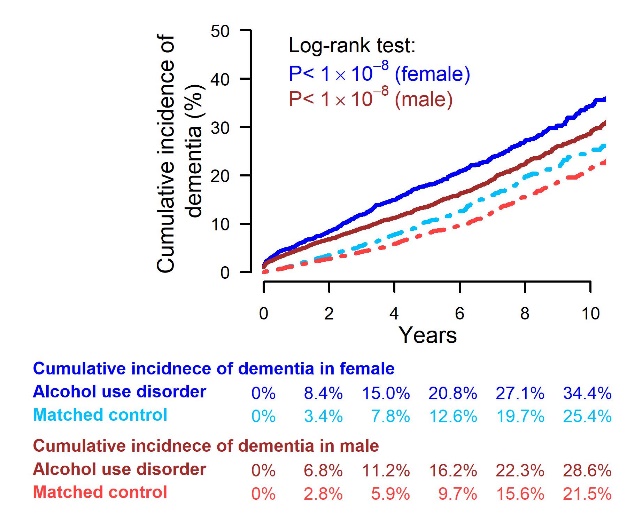 |
| **C.** Black subpopulation | **D.** Asian subpopulation |
| 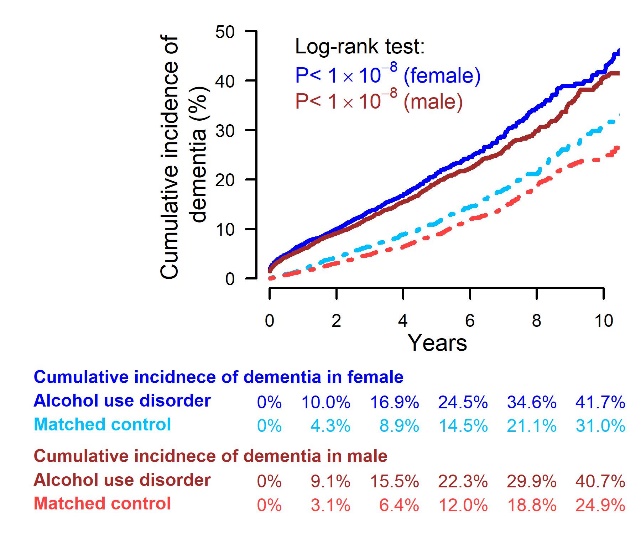 | 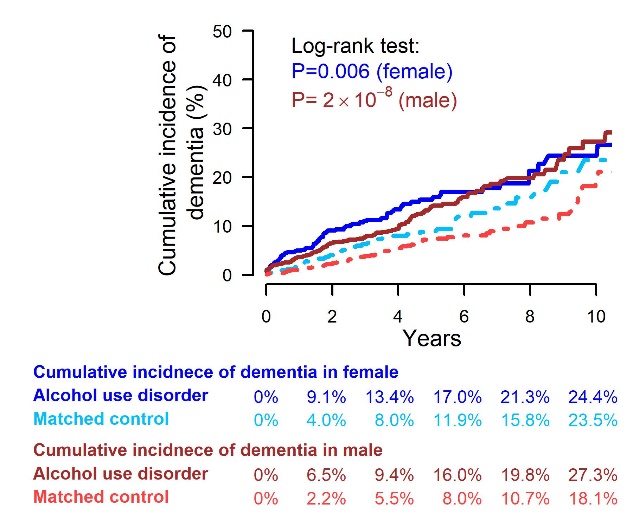 |

**Figure S2.** Cumulative incidence of dementia for alcohol use disorder patients and matched controls; **A.** All Races (female N=90,486, male N=167,878); **B.** White subpopulation (female N=68,460, male N=119,808); **C.** Hispanic subpopulation (female N=7,210, male N=20,442); **D.** Black subpopulation (female N=8,294, male N=15,234); **E.** Asian subpopulation (female N=1,352, male N=3,304).

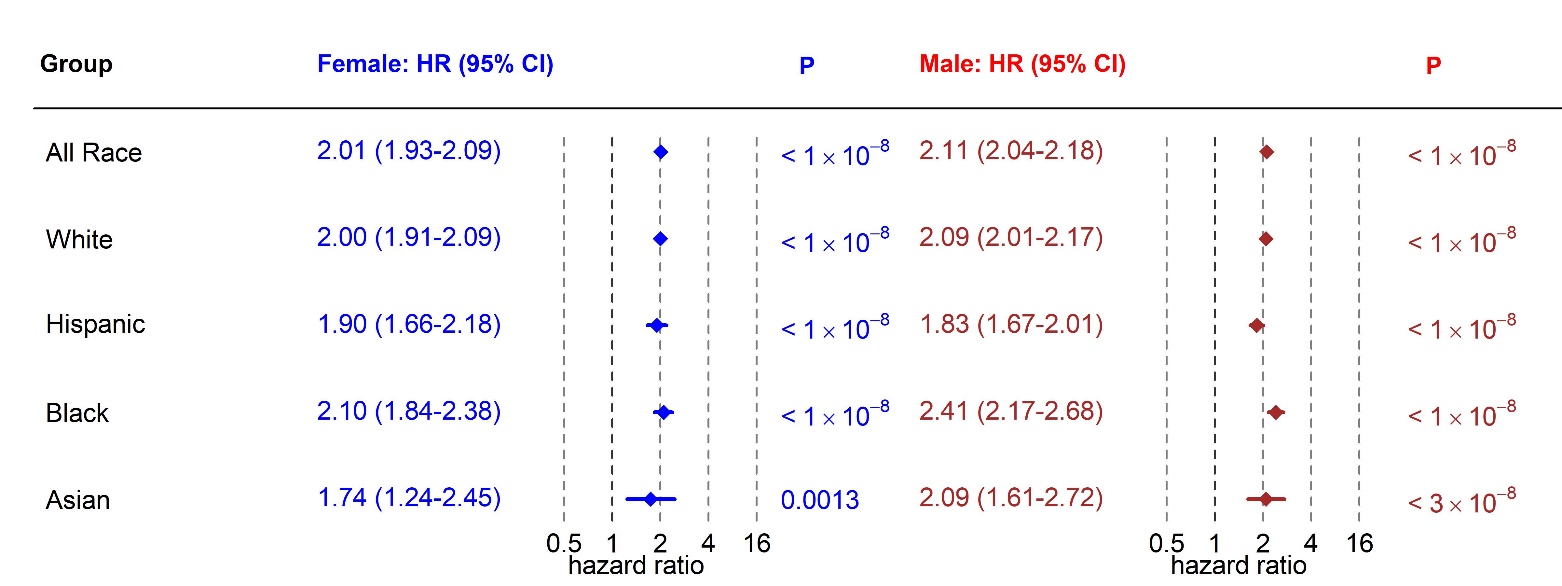

**Figure S3** Association of alcohol use disorders with dementia in multivariate Cox analyses (N=258,364).

| 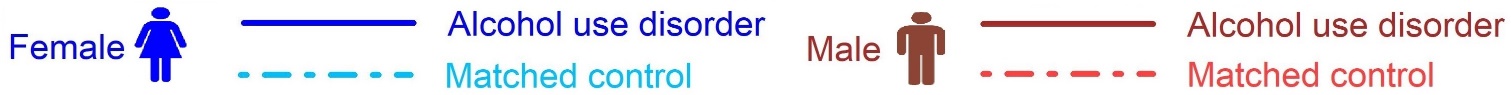 | |
| --- | --- |
| **A.** All Races | |
| 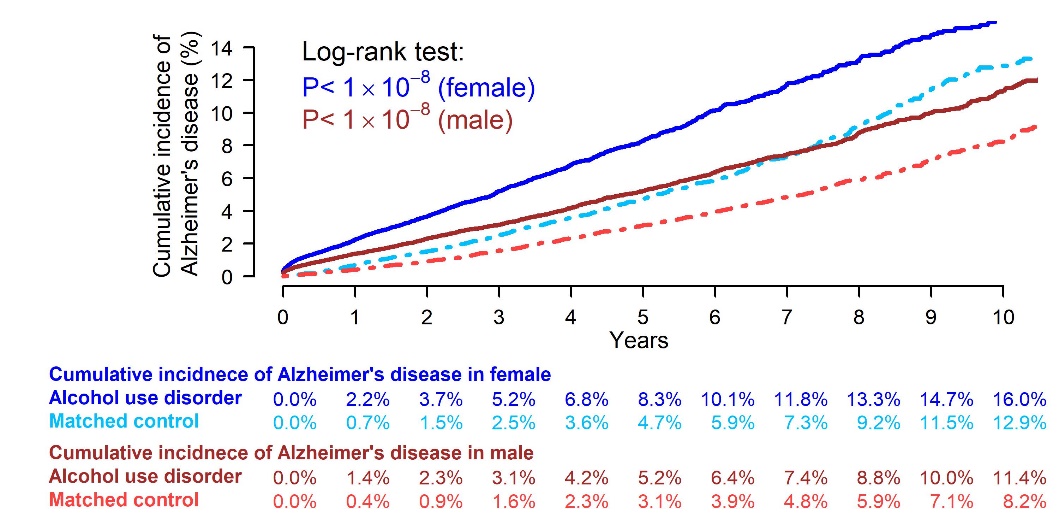 | |
| **B.** White subpopulation | **C.** Hispanic subpopulation |
| 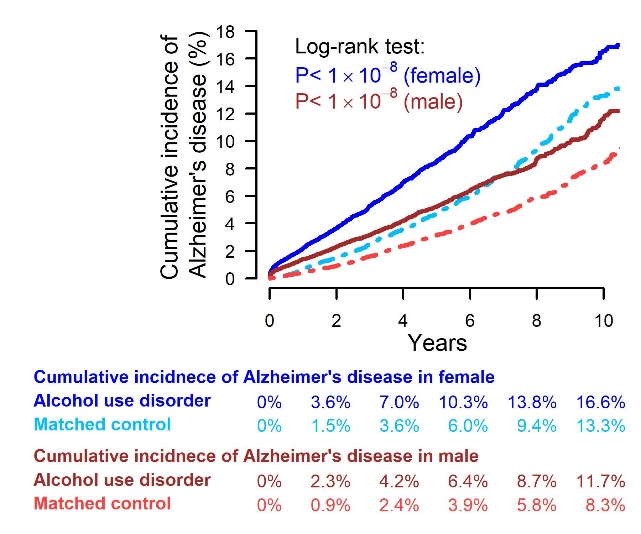 | 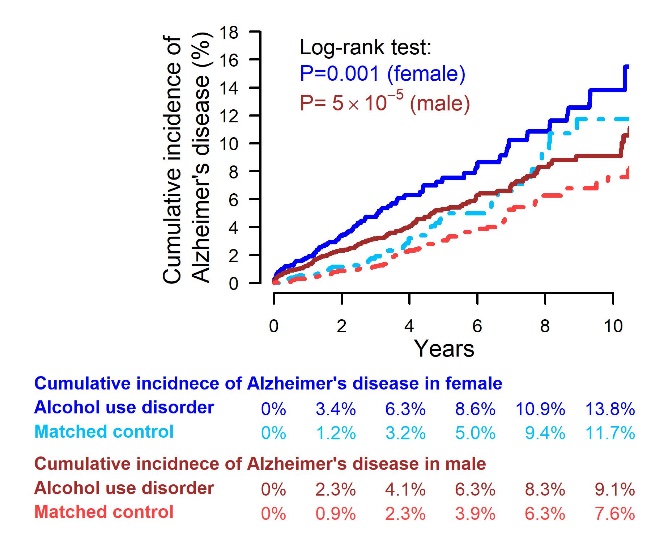 |
| **C.** Black subpopulation | **D.** Asian subpopulation |
| 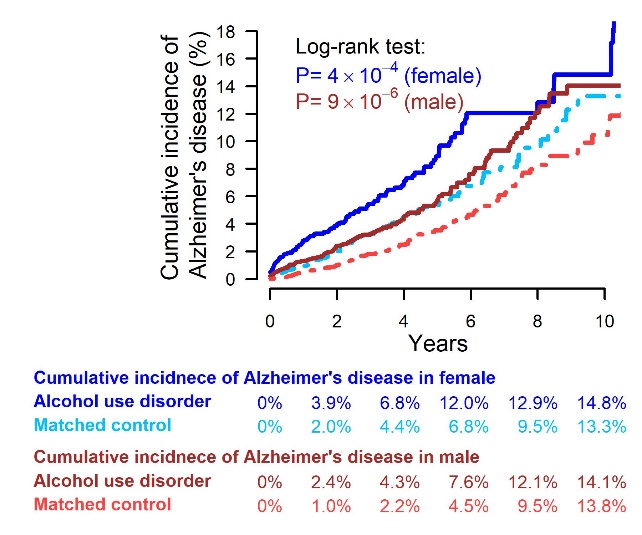 | 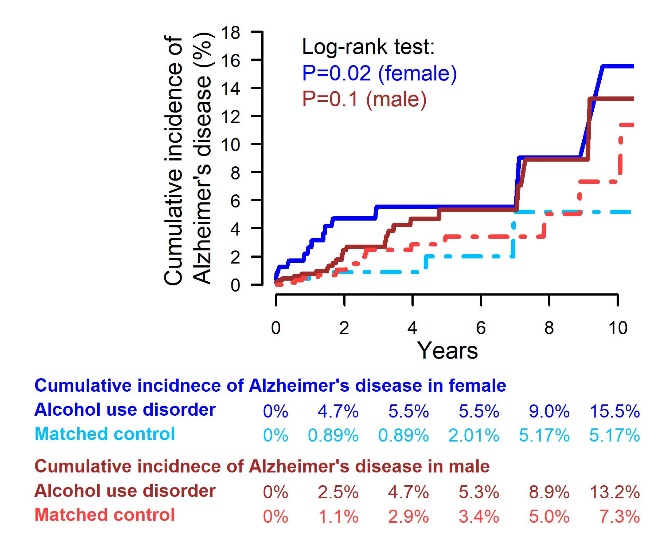 |

**Figure S4.** Cumulative incidence of Alzheimer’s disease for patients with alcohol abuse and matched controls; **A.** All Races (female N=42,164, male N=77,884); **B.** White subpopulation (female N=32,418, male N=56,468); **C.** Hispanic subpopulation (female N=2,768, male N=7,574); **D.** Black subpopulation (female N=4,194, male N=8,408); **E.** Asian subpopulation (female N=476, male N=1,364).

| 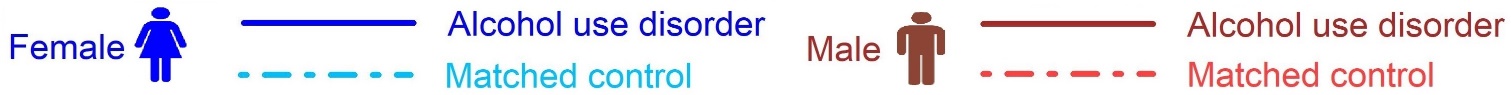 | |
| --- | --- |
| **A.** All Races | |
| 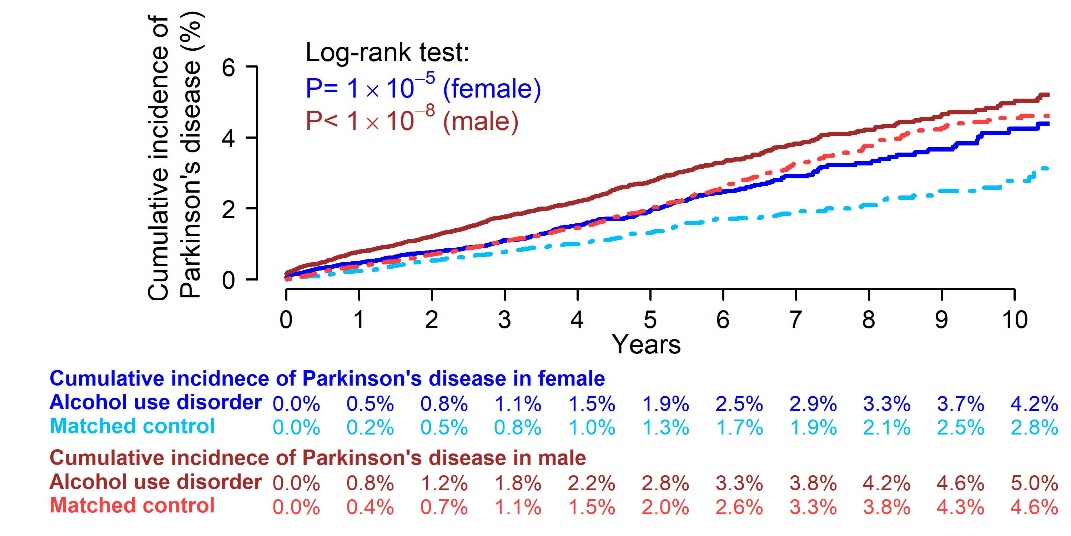 | |
| **B.** White subpopulation | **C.** Hispanic subpopulation |
| 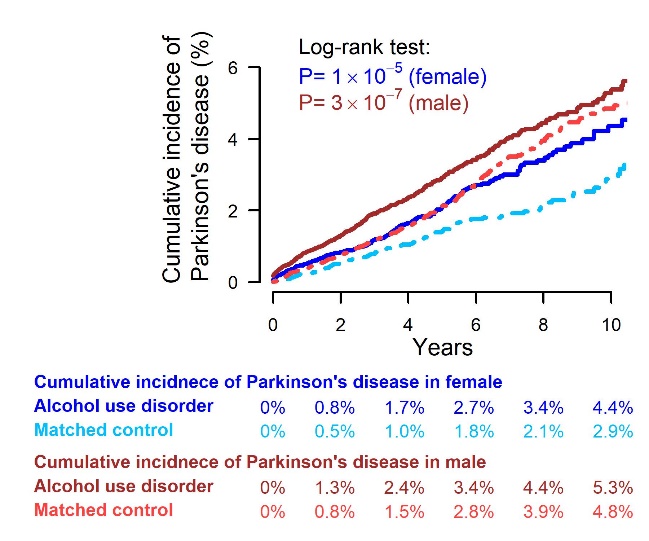 | 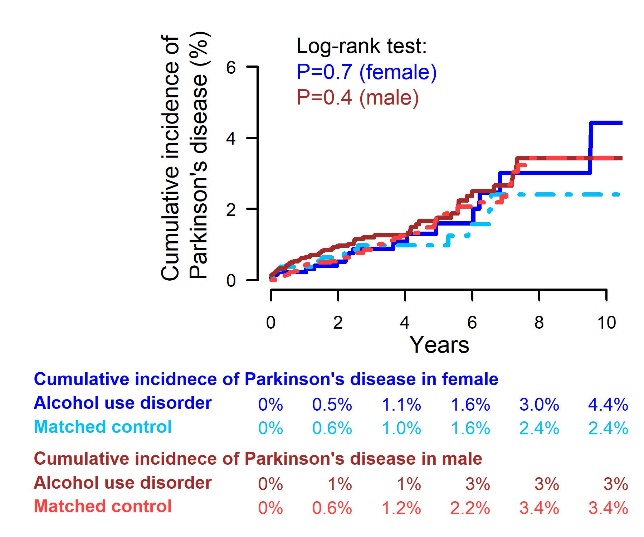 |
| **C.** Black subpopulation | **D.** Asian subpopulation |
| 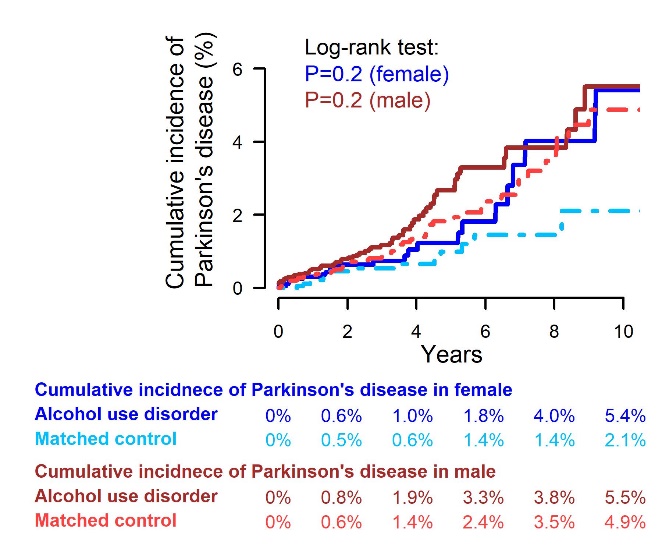 | 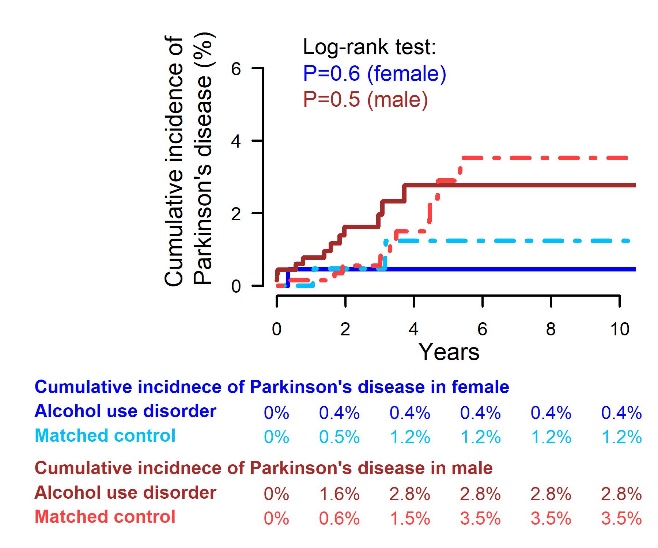 |

**Figure S5.** Cumulative incidence of Parkinson's disease for patients with alcohol abuse and matched controls; **A.** All Races (female N=42,164, male N=77,884); **B.** White subpopulation (female N=32,418, male N=56,468); **C.** Hispanic subpopulation (female N=2,768, male N=7,574); **D.** Black subpopulation (female N=4,194, male N=8,408); **E.** Asian subpopulation (female N=476, male N=1,364).

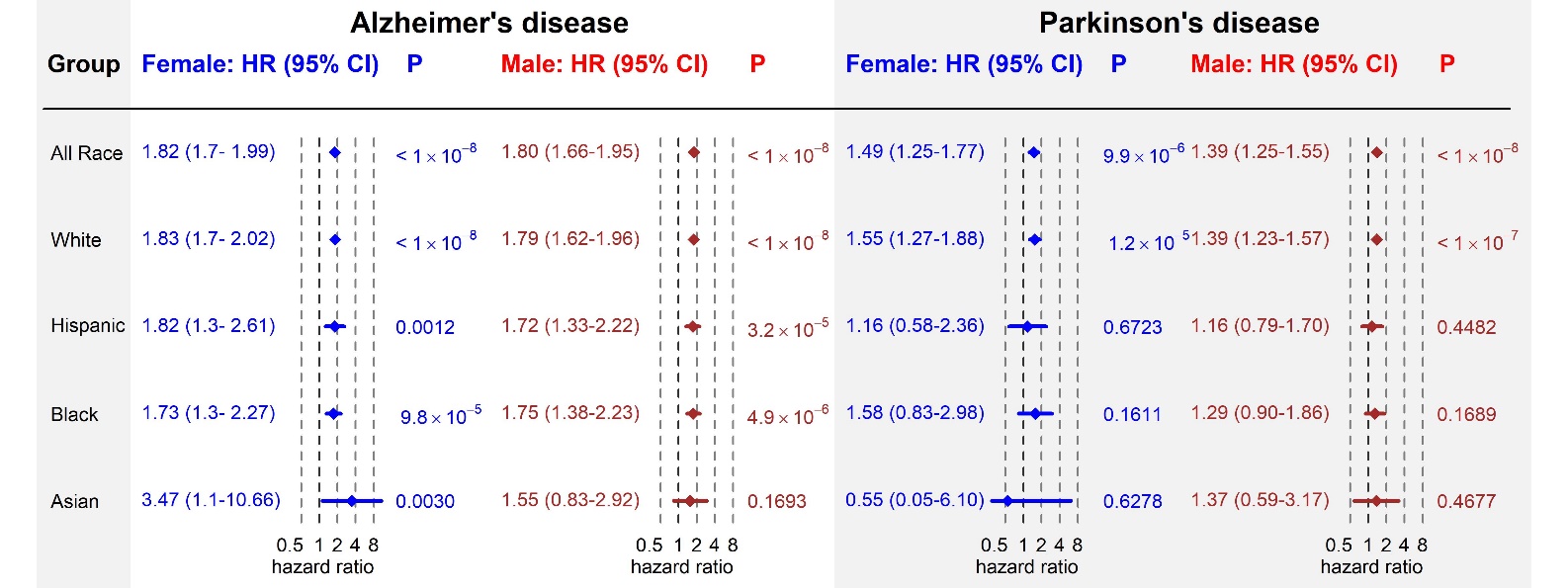

**Figure S6.** Association of alcohol abuse with Alzheimer’s disease and Parkinson's disease in multivariate Cox analyses (N= 120,048).

| 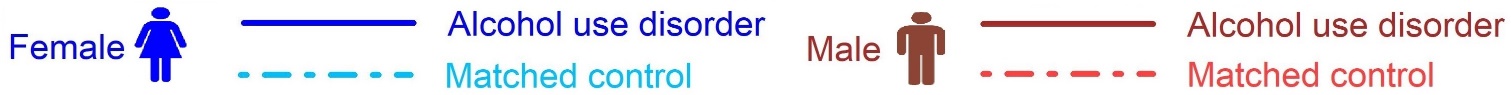 | |
| --- | --- |
| **A.** All Races | |
| 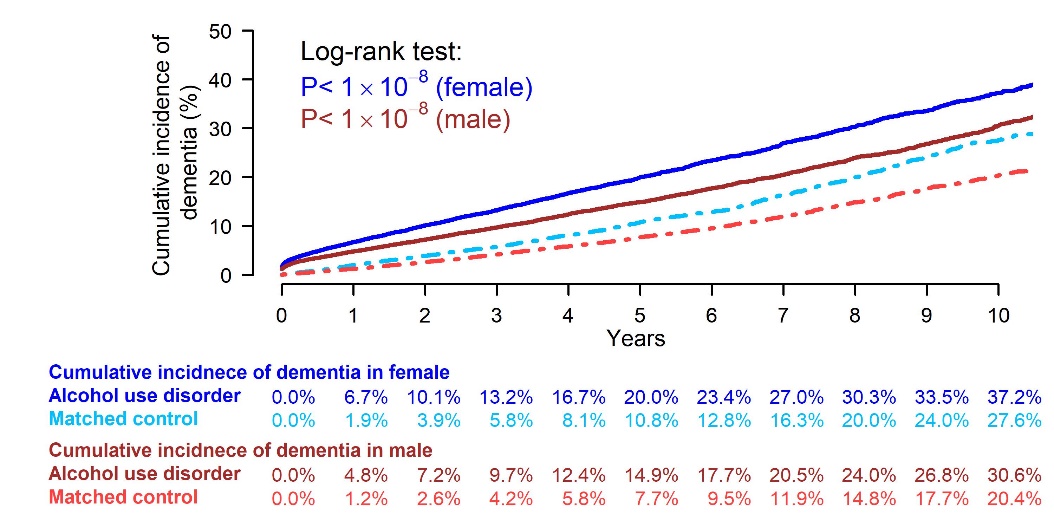 | |
| **B.** White subpopulation | **C.** Hispanic subpopulation |
| 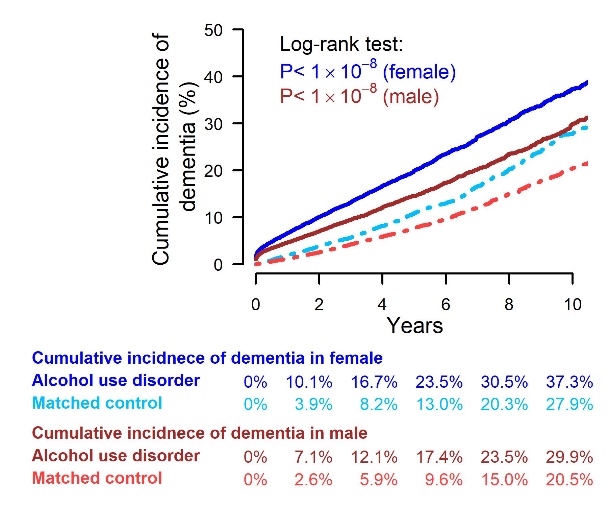 | 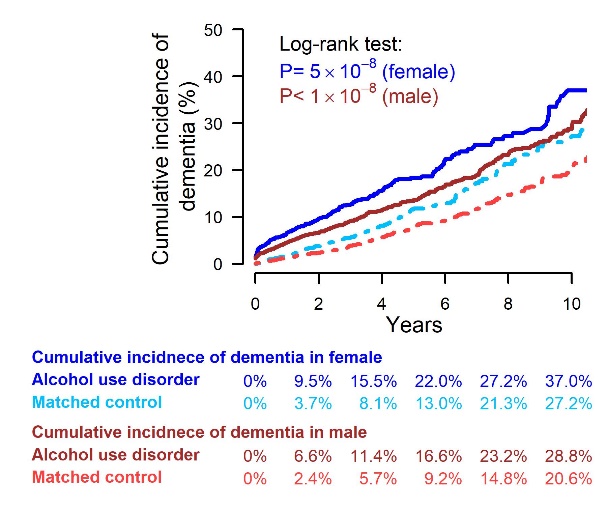 |
| **C.** Black subpopulation | **D.** Asian subpopulation |
| 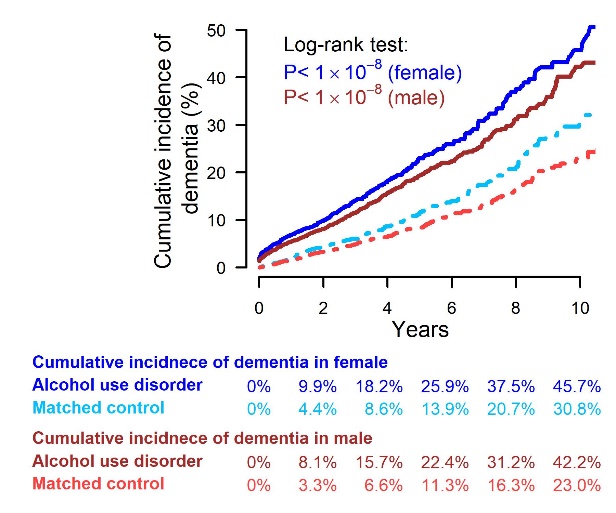 | 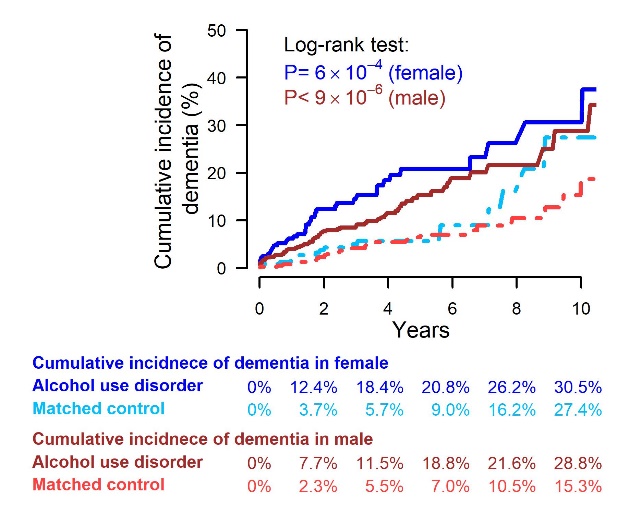 |

**Figure S7.** Cumulative incidence of dementia for alcohol abuse patients and matched controls; **A.** All Races (female N=42,164, male N=77,884); **B.** White subpopulation (female N=32,418, male N=56,468); **C.** Hispanic subpopulation (female N=2,768, male N=7,574); **D.** Black subpopulation (female N=4,194, male N=8,408); **E.** Asian subpopulation (female N=476, male N=1,364).

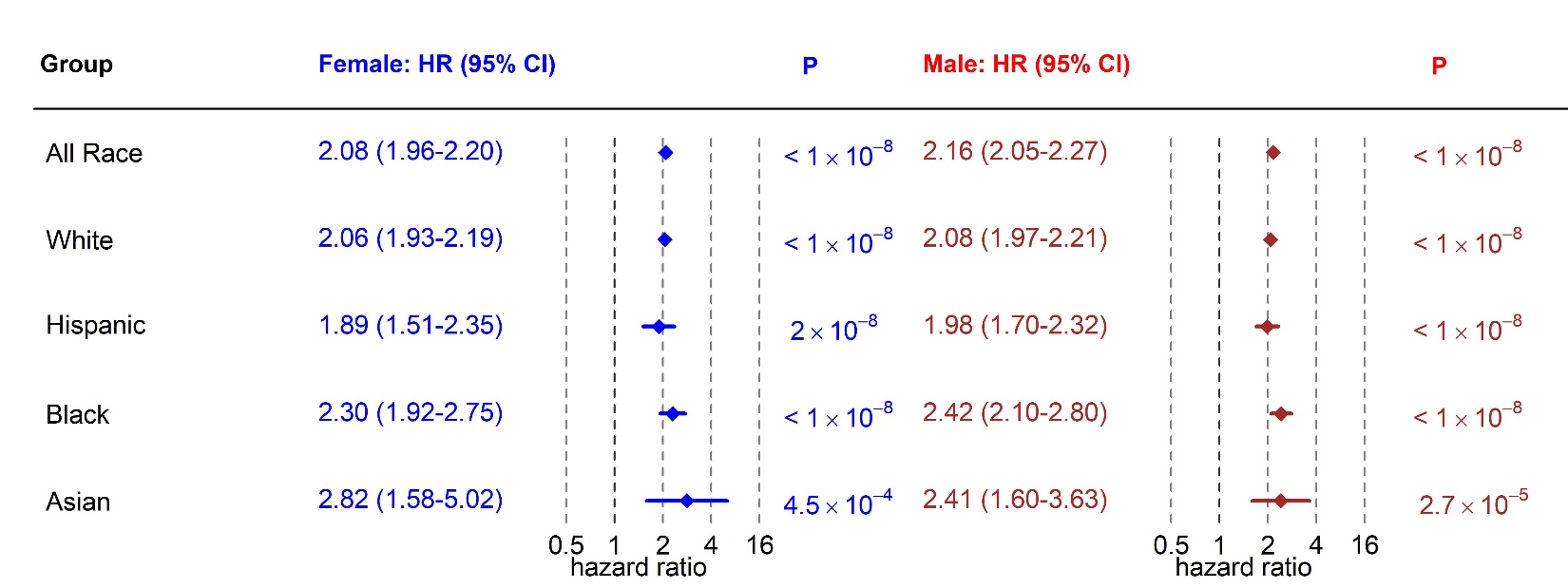

**Figure S8.** Association of alcohol abuse with dementia in multivariate Cox analyses (N=120,048).

**Table S1.** Demographics of race-specific subpopulations of patients with alcohol use disorder.

|  | Female | | | | Male | | | |
| --- | --- | --- | --- | --- | --- | --- | --- | --- |
|  | Alcohol use disorder | | Matched Control | | Alcohol use disorder | | Matched Control | |
| ***White subpopulation*** | | | | | | | | |
| **Age group** | | | | | | | | |
| 65-75 | 22,722 | 66.40% | 22,722 | 66.40% | 43,038 | 71.80% | 43,038 | 71.80% |
| 76-100 | 11,508 | 33.60% | 11,508 | 33.60% | 16,866 | 28.20% | 16,866 | 28.20% |
| **Comorbidity** |  |  |  |  |  |  |  |  |
| Cerebrovascular disease | 7,292 | 21.30% | 7,292 | 21.30% | 12,373 | 20.70% | 12,373 | 20.70% |
| Depression | 13,041 | 38.10% | 13,041 | 38.10% | 11,610 | 19.40% | 11,610 | 19.40% |
| Diabetes | 7,831 | 22.90% | 7,831 | 22.90% | 17,807 | 29.70% | 17,807 | 29.70% |
| Fall | 6,861 | 20.00% | 6,861 | 20.00% | 6,451 | 10.80% | 6,451 | 10.80% |
| Heart disease | 5,996 | 17.50% | 5,996 | 17.50% | 13,902 | 23.20% | 13,902 | 23.20% |
| Hypertension | 26,118 | 76.30% | 26,118 | 76.30% | 47,110 | 78.60% | 47,110 | 78.60% |
| Liver disease | 5,338 | 15.60% | 5,338 | 15.60% | 8,042 | 13.40% | 8,042 | 13.40% |
| Chronic pulmonary disease | 13,909 | 40.60% | 13,909 | 40.60% | 21,712 | 36.20% | 21,712 | 36.20% |
| Renal disease | 5,505 | 16.10% | 5,505 | 16.10% | 9,888 | 16.50% | 9,888 | 16.50% |
| Traumatic brain injury | 621 | 1.80% | 621 | 1.80% | 556 | 0.90% | 556 | 0.90% |
| ***Hispanic subpopulation*** | | | | | | | | |
| **Age group** | | | | | | | | |
| 65-75 | 2,418 | 67.10% | 2,418 | 67.10% | 7,331 | 71.70% | 7,331 | 71.70% |
| 76-100 | 1,187 | 32.90% | 1,187 | 32.90% | 2,890 | 28.30% | 2,890 | 28.30% |
| **Comorbidity** |  |  |  |  |  |  |  |  |
| Cerebrovascular disease | 686 | 19.00% | 686 | 19.00% | 1,593 | 15.60% | 1,593 | 15.60% |
| Depression | 1,256 | 34.80% | 1,256 | 34.80% | 1,466 | 14.30% | 1,466 | 14.30% |
| Diabetes | 1,522 | 42.20% | 1,522 | 42.20% | 4,655 | 45.50% | 4,655 | 45.50% |
| Fall | 591 | 16.40% | 591 | 16.40% | 737 | 7.20% | 737 | 7.20% |
| Heart disease | 665 | 18.40% | 665 | 18.40% | 2,034 | 19.90% | 2,034 | 19.90% |
| Hypertension | 2,924 | 81.10% | 2,924 | 81.10% | 7,829 | 76.60% | 7,829 | 76.60% |
| Liver disease | 873 | 24.20% | 873 | 24.20% | 1,622 | 15.90% | 1,622 | 15.90% |
| Chronic pulmonary disease | 1,328 | 36.80% | 1,328 | 36.80% | 2,884 | 28.20% | 2,884 | 28.20% |
| Renal disease | 834 | 23.10% | 834 | 23.10% | 2,331 | 22.80% | 2,331 | 22.80% |
| Traumatic brain injury | 36 | 1.00% | 36 | 1.00% | 31 | 0.30% | 31 | 0.30% |
| ***Black subpopulation*** | | | | | | | | |
| **Age group** | | | | | | | | |
| 65-75 | 3,101 | 74.80% | 3,101 | 74.80% | 6,065 | 79.60% | 6,065 | 79.60% |
| 76-100 | 1,046 | 25.20% | 1,046 | 25.20% | 1,552 | 20.40% | 1,552 | 20.40% |
| **Comorbidity** |  |  |  |  |  |  |  |  |
| Cerebrovascular disease | 828 | 20.00% | 828 | 20.00% | 1,356 | 17.80% | 1,356 | 17.80% |
| Depression | 1,313 | 31.70% | 1,313 | 31.70% | 900 | 11.80% | 900 | 11.80% |
| Diabetes | 1,527 | 36.80% | 1,527 | 36.80% | 2,723 | 35.70% | 2,723 | 35.70% |
| Fall | 622 | 15.00% | 622 | 15.00% | 519 | 6.80% | 519 | 6.80% |
| Heart disease | 881 | 21.20% | 881 | 21.20% | 1,789 | 23.50% | 1,789 | 23.50% |
| Hypertension | 3,556 | 85.70% | 3,556 | 85.70% | 6,212 | 81.60% | 6,212 | 81.60% |
| Liver disease | 634 | 15.30% | 634 | 15.30% | 843 | 11.10% | 843 | 11.10% |
| Chronic pulmonary disease | 1,745 | 42.10% | 1,745 | 42.10% | 2,570 | 33.70% | 2,570 | 33.70% |
| Renal disease | 804 | 19.40% | 804 | 19.40% | 1,486 | 19.50% | 1,486 | 19.50% |
| Traumatic brain injury | 22 | 0.50% | 22 | 0.50% | 22 | 0.30% | 22 | 0.30% |
| ***Asian subpopulation*** | | | | | | | | |
| **Age group** | | | | | | | | |
| 65-75 | 456 | 67.50% | 456 | 67.50% | 1,187 | 71.90% | 1,187 | 71.90% |
| 76-100 | 220 | 32.50% | 220 | 32.50% | 465 | 28.10% | 465 | 28.10% |
| **Comorbidity** |  |  |  |  |  |  |  |  |
| Cerebrovascular disease | 105 | 15.50% | 105 | 15.50% | 290 | 17.60% | 290 | 17.60% |
| Depression | 163 | 24.10% | 163 | 24.10% | 114 | 6.90% | 114 | 6.90% |
| Diabetes | 231 | 34.20% | 231 | 34.20% | 722 | 43.70% | 722 | 43.70% |
| Fall | 72 | 10.70% | 72 | 10.70% | 68 | 4.10% | 68 | 4.10% |
| Heart disease | 85 | 12.60% | 85 | 12.60% | 236 | 14.30% | 236 | 14.30% |
| Hypertension | 539 | 79.70% | 539 | 79.70% | 1,329 | 80.40% | 1,329 | 80.40% |
| Liver disease | 161 | 23.80% | 161 | 23.80% | 276 | 16.70% | 276 | 16.70% |
| Chronic pulmonary disease | 201 | 29.70% | 201 | 29.70% | 470 | 28.50% | 470 | 28.50% |
| Renal disease | 100 | 14.80% | 100 | 14.80% | 320 | 19.40% | 320 | 19.40% |
| Traumatic brain injury | 1 | 0.10% | 1 | 0.10% | 5 | 0.30% | 5 | 0.30% |

**Table S2.** Results from multivariate Cox analysis: association with Alzheimer’s disease (N=247,310).

|  | **Female** | | **male** | |
| --- | --- | --- | --- | --- |
| **Variable** | HR (95% CI) | P | HR (95% CI) | P |
| **All Races** | | | | |
| Alcohol use disorder | 1.78 (1.68-1.90) | <0.0001 | 1.80 (1.71-1.91) | <0.0001 |
| **Age (referent = 65-74)** |  |  |  |  |
| 76-100 | 3.83 (3.59-4.09) | <0.0001 | 3.53 (3.34-3.73) | <0.0001 |
| **Race (referent = White)** |  |  |  |  |
| Hispanic | 0.96 (0.86-1.07) | 0.4970 | 0.94 (0.87-1.02) | 0.1459 |
| Black | 1.25 (1.13-1.39) | <0.0001 | 1.33 (1.21-1.46) | <0.0001 |
| Asian | 0.76 (0.57-1.00) | 0.0537 | 0.74 (0.59-0.93) | 0.0097 |
| Unknown | 0.89 (0.76-1.03) | 0.1060 | 0.79 (0.68-0.91) | 0.0013 |
| **Comorbidity** |  |  |  |  |
| Cerebrovascular disease | 1.26 (1.17-1.36) | <0.0001 | 1.40 (1.31-1.49) | <0.0001 |
| Depression | 1.32 (1.24-1.41) | <0.0001 | 1.41 (1.32-1.52) | <0.0001 |
| Diabetes | 1.02 (0.95-1.10) | 0.6326 | 1.12 (1.06-1.19) | 0.0002 |
| Fall | 1.27 (1.18-1.37) | <0.0001 | 1.32 (1.21-1.44) | <0.0001 |
| Heart disease | 1.05 (0.97-1.14) | 0.2525 | 1.00 (0.94-1.07) | 0.9658 |
| Hypertension | 1.00 (0.92-1.08) | 0.9850 | 0.85 (0.79-0.91) | <0.0001 |
| Liver disease | 0.77 (0.70-0.86) | <0.0001 | 0.73 (0.66-0.81) | <0.0001 |
| Chronic pulmonary disease | 0.86 (0.81-0.92) | <0.0001 | 0.92 (0.87-0.98) | 0.0076 |
| Renal disease | 0.89 (0.82-0.97) | 0.0057 | 1.03 (0.95-1.10) | 0.4772 |
| Traumatic brain injury | 1.15 (0.93-1.44) | 0.1954 | 1.12 (0.84-1.49) | 0.4252 |
| **White subpopulation** | | | | |
| Alcohol use disorder | 1.78 (1.66-1.91) | <0.0001 | 1.81 (1.70-1.93) | <0.0001 |
| **Age (referent = 65-74)** |  |  |  |  |
| 76-100 | 3.86 (3.58-4.15) | <0.0001 | 3.51 (3.29-3.75) | <0.0001 |
| **Comorbidity** |  |  |  |  |
| Cerebrovascular disease | 1.27 (1.18-1.38) | <0.0001 | 1.42 (1.32-1.53) | <0.0001 |
| Depression | 1.33 (1.24-1.43) | <0.0001 | 1.41 (1.30-1.52) | <0.0001 |
| Diabetes | 1.03 (0.95-1.12) | 0.4604 | 1.13 (1.06-1.22) | 0.0006 |
| Fall | 1.33 (1.22-1.44) | <0.0001 | 1.32 (1.20-1.45) | <0.0001 |
| Heart disease | 1.04 (0.95-1.14) | 0.4079 | 0.99 (0.91-1.07) | 0.7696 |
| Hypertension | 1.00 (0.92-1.09) | 0.9922 | 0.84 (0.77-0.91) | <0.0001 |
| Liver disease | 0.78 (0.70-0.88) | <0.0001 | 0.75 (0.67-0.84) | <0.0001 |
| Chronic pulmonary disease | 0.84 (0.78-0.90) | <0.0001 | 0.94 (0.87-1.00) | 0.0651 |
| Renal disease | 0.91 (0.82-1.00) | 0.0426 | 1.02 (0.94-1.12) | 0.5877 |
| Traumatic brain injury | 1.18 (0.94-1.48) | 0.1474 | 0.97 (0.70-1.34) | 0.8505 |
| **Hispanic subpopulation** | | | | |
| Alcohol use disorder | 1.58 (1.28-1.95) | <0.0001 | 1.59 (1.37-1.84) | <0.0001 |
| **Age (referent = 65-74)** |  |  |  |  |
| 76-100 | 3.15 (2.53-3.92) | <0.0001 | 3.53 (3.03-4.11) | <0.0001 |
| **Comorbidity** |  |  |  |  |
| Cerebrovascular disease | 1.03 (0.78-1.36) | 0.8420 | 1.44 (1.19-1.75) | 0.0002 |
| Depression | 1.06 (0.84-1.34) | 0.6415 | 1.62 (1.32-2.00) | <0.0001 |
| Diabetes | 0.85 (0.67-1.07) | 0.1608 | 1.02 (0.86-1.21) | 0.7898 |
| Fall | 1.09 (0.81-1.47) | 0.5554 | 1.34 (1.02-1.75) | 0.0346 |
| Heart disease | 0.90 (0.67-1.22) | 0.5087 | 1.06 (0.87-1.29) | 0.5835 |
| Hypertension | 1.06 (0.80-1.41) | 0.6713 | 0.85 (0.70-1.03) | 0.0915 |
| Liver disease | 0.94 (0.72-1.23) | 0.6589 | 0.74 (0.57-0.96) | 0.0213 |
| Chronic pulmonary disease | 1.04 (0.83-1.31) | 0.7246 | 0.83 (0.69-0.99) | 0.0372 |
| Renal disease | 1.01 (0.77-1.32) | 0.9436 | 0.96 (0.80-1.17) | 0.7078 |
| Traumatic brain injury |  |  | 1.09 (0.27-4.46) | 0.9030 |
| **Black subpopulation** | | | | |
| Alcohol use disorder | 1.98 (1.61-2.44) | <0.0001 | 1.95 (1.63-2.33) | <0.0001 |
| **Age (referent = 65-74)** |  |  |  |  |
| 76-100 | 3.67 (2.98-4.52) | <0.0001 | 3.24 (2.71-3.88) | <0.0001 |
| **Comorbidity** |  |  |  |  |
| Cerebrovascular disease | 1.33 (1.05-1.69) | 0.0187 | 1.28 (1.03-1.60) | 0.0271 |
| Depression | 1.39 (1.11-1.74) | 0.0037 | 1.25 (0.94-1.64) | 0.1217 |
| Diabetes | 0.98 (0.78-1.24) | 0.8871 | 1.10 (0.90-1.33) | 0.3653 |
| Fall | 1.03 (0.77-1.38) | 0.8427 | 1.35 (0.96-1.88) | 0.0811 |
| Heart disease | 1.30 (1.01-1.67) | 0.0439 | 1.06 (0.84-1.33) | 0.6261 |
| Hypertension | 1.00 (0.74-1.35) | 0.9950 | 0.97 (0.76-1.23) | 0.7811 |
| Liver disease | 0.57 (0.38-0.87) | 0.0094 | 0.63 (0.43-0.91) | 0.0151 |
| Chronic pulmonary disease | 0.94 (0.75-1.17) | 0.5638 | 0.84 (0.68-1.03) | 0.0937 |
| Renal disease | 0.72 (0.54-0.96) | 0.0241 | 1.18 (0.93-1.49) | 0.1633 |
| Traumatic brain injury |  |  | 2.95 (1.17-7.43) | 0.0214 |
| **Asian subpopulation** | | | | |
| Alcohol use disorder | 2.49 (1.35-4.57) | 0.0034 | 1.70 (1.07-2.69) | 0.0242 |
| **Age (referent = 65-74)** |  |  |  |  |
| 76-100 | 3.50 (1.92-6.38) | <0.0001 | 3.07 (1.91-4.92) | <0.0001 |
| **Comorbidity** |  |  |  |  |
| Cerebrovascular disease | 1.49 (0.68-3.26) | 0.3155 | 0.76 (0.40-1.43) | 0.3913 |
| Depression | 1.32 (0.68-2.58) | 0.4119 | 3.24 (1.77-5.95) | 1.00E-04 |
| Diabetes | 0.46 (0.21-1.00) | 0.0507 | 1.32 (0.81-2.16) | 0.2677 |
| Fall | 0.29 (0.07-1.22) | 0.0906 | 1.88 (0.74-4.77) | 0.1856 |
| Heart disease | 2.25 (0.96-5.26) | 0.0605 | 0.83 (0.42-1.64) | 0.5912 |
| Hypertension | 0.82 (0.40-1.67) | 0.5899 | 0.96 (0.50-1.84) | 0.9022 |
| Liver disease | 0.58 (0.24-1.38) | 0.2155 | 0.67 (0.32-1.40) | 0.2837 |
| Chronic pulmonary disease | 0.82 (0.42-1.60) | 0.5555 | 0.94 (0.56-1.59) | 0.8187 |

**Table S3.** Results from multivariate Cox analysis: association with Parkinson's disease (N=247,310).

|  | **Female** | | **male** | |
| --- | --- | --- | --- | --- |
| **Variable** | HR (95% CI) | P | HR (95% CI) | P |
| **All Races** | | | | |
| Alcohol use disorder | 1.49 (1.32-1.68) | <0.0001 | 1.42 (1.32-1.52) | <0.0001 |
| **Age (referent = 65-74)** |  |  |  |  |
| 76-100 | 1.43 (1.26-1.61) | <0.0001 | 1.66 (1.54-1.79) | <0.0001 |
| **Race (referent = White)** |  |  |  |  |
| Hispanic | 0.78 (0.62-0.98) | 0.0325 | 0.79 (0.70-0.88) | <0.0001 |
| Black | 0.90 (0.72-1.13) | 0.3632 | 0.86 (0.74-0.99) | 0.0348 |
| Asian | 0.76 (0.44-1.31) | 0.3183 | 0.84 (0.63-1.11) | 0.2209 |
| Unknown | 0.98 (0.75-1.28) | 0.8625 | 0.9 (0.75-1.07) | 0.2292 |
| **Comorbidity** |  |  |  |  |
| Cerebrovascular disease | 1.32 (1.15-1.52) | 0.0001 | 1.32 (1.21-1.45) | <0.0001 |
| Depression | 1.72 (1.53-1.94) | <0.0001 | 1.83 (1.68-2.00) | <0.0001 |
| Diabetes | 1.15 (1.00-1.32) | 0.0543 | 1.20 (1.10-1.30) | <0.0001 |
| Fall | 1.34 (1.15-1.55) | 0.0001 | 1.31 (1.16-1.47) | <0.0001 |
| Heart disease | 1.12 (0.95-1.31) | 0.1653 | 0.93 (0.85-1.03) | 0.1482 |
| Hypertension | 0.95 (0.82-1.10) | 0.4924 | 0.92 (0.84-1.01) | 0.0846 |
| Liver disease | 1.02 (0.86-1.21) | 0.8174 | 0.82 (0.72-0.92) | 0.0013 |
| Chronic pulmonary disease | 0.92 (0.81-1.05) | 0.2043 | 0.92 (0.85-0.99) | 0.0332 |
| Renal disease | 0.96 (0.82-1.14) | 0.6595 | 1.00 (0.90-1.10) | 0.9705 |
| Traumatic brain injury | 1.32 (0.89-1.97) | 0.1718 | 1.49 (1.07-2.09) | 0.0198 |
| **White subpopulation** | | | | |
| Alcohol use disorder | 1.55 (1.36-1.77) | <0.0001 | 1.45 (1.33-1.57) | <0.0001 |
| **Age (referent = 65-74)** |  |  |  |  |
| 76-100 | 1.37 (1.19-1.58) | <0.0001 | 1.58 (1.45-1.73) | <0.0001 |
| **Comorbidity** |  |  |  |  |
| Cerebrovascular disease | 1.31 (1.12-1.54) | 7.00E-04 | 1.28 (1.16-1.42) | <0.0001 |
| Depression | 1.64 (1.43-1.87) | <0.0001 | 1.83 (1.66-2.01) | <0.0001 |
| Diabetes | 1.22 (1.04-1.42) | 0.0149 | 1.27 (1.15-1.39) | <0.0001 |
| Fall | 1.34 (1.14-1.58) | 0.0003 | 1.34 (1.18-1.52) | <0.0001 |
| Heart disease | 1.14 (0.95-1.36) | 0.1534 | 0.94 (0.85-1.05) | 0.2910 |
| Hypertension | 0.91 (0.78-1.08) | 0.2927 | 0.94 (0.85-1.05) | 0.2844 |
| Liver disease | 1.01 (0.83-1.21) | 0.9588 | 0.77 (0.67-0.89) | 0.0003 |
| Chronic pulmonary disease | 0.95 (0.82-1.09) | 0.4579 | 0.90 (0.82-0.99) | 0.0242 |
| Renal disease | 0.94 (0.78-1.14) | 0.5504 | 0.99 (0.88-1.11) | 0.8731 |
| Traumatic brain injury | 1.41 (0.93-2.11) | 0.1024 | 1.48 (1.04-2.10) | 0.0307 |
| **Hispanic subpopulation** | | | | |
| Alcohol use disorder | 1.12 (0.73-1.74) | 0.6017 | 1.22 (0.98-1.51) | 0.0715 |
| **Age (referent = 65-74)** |  |  |  |  |
| 76-100 | 1.69 (1.07-2.67) | 0.0241 | 2.01 (1.61-2.50) | <0.0001 |
| **Comorbidity** |  |  |  |  |
| Cerebrovascular disease | 0.88 (0.48-1.59) | 0.6614 | 1.33 (0.99-1.77) | 0.0569 |
| Depression | 1.70 (1.07-2.68) | 0.0237 | 1.94 (1.46-2.57) | <0.0001 |
| Diabetes | 1.02 (0.63-1.66) | 0.9315 | 0.97 (0.76-1.24) | 0.8016 |
| Fall | 0.98 (0.52-1.85) | 0.9557 | 1.06 (0.68-1.65) | 0.8031 |
| Heart disease | 1.19 (0.66-2.16) | 0.5547 | 0.96 (0.71-1.30) | 0.8032 |
| Hypertension | 0.91 (0.50-1.67) | 0.7636 | 0.88 (0.67- 1.16) | 0.3807 |
| Liver disease | 1.17 (0.69-1.96) | 0.5598 | 1.23 (0.90-1.67) | 0.1871 |
| Chronic pulmonary disease | 1.16 (0.73-1.84) | 0.5411 | 0.83 (0.64-1.08) | 0.1725 |
| Renal disease | 0.99 (0.57-1.74) | 0.9855 | 1.04 (0.78-1.37) | 0.8032 |
| Traumatic brain injury |  |  | 3.39 (0.81-14.1) | 0.0942 |
| **Black subpopulation** | | | | |
| Alcohol use disorder | 1.36 (0.88-2.10) | 0.1673 | 1.32 (1.01-1.74) | 0.0459 |
| **Age (referent = 65-74)** |  |  |  |  |
| 76-100 | 1.68 (1.06-2.68) | 0.0286 | 1.96 (1.46-2.64) | <0.0001 |
| **Comorbidity** |  |  |  |  |
| Cerebrovascular disease | 1.57 (0.95-2.59) | 0.0817 | 1.63 (1.17-2.28) | 0.0042 |
| Depression | 2.73 (1.74-4.26) | <0.0001 | 1.72 (1.18-2.52) | 0.0048 |
| Diabetes | 0.80 (0.49-1.32) | 0.3834 | 1.00 (0.73-1.36) | 0.9952 |
| Fall | 1.11 (0.60-2.05) | 0.7292 | 1.12 (0.64-1.97) | 0.6924 |
| Heart disease | 1.01 (0.57-1.79) | 0.9694 | 0.84 (0.58-1.22) | 0.3547 |
| Hypertension | 1.20 (0.59-2.42) | 0.6165 | 1.02 (0.69-1.49) | 0.9338 |
| Liver disease | 1.13 (0.57-2.21) | 0.7297 | 0.72 (0.42-1.24) | 0.2385 |
| Chronic pulmonary disease | 0.80 (0.50-1.29) | 0.3656 | 1.03 (0.76-1.41) | 0.8310 |
| Renal disease | 1.40 (0.82-2.40) | 0.2153 | 0.98 (0.66-1.44) | 0.8982 |
| Traumatic brain injury |  |  | 1.49 (0.20-11.23) | 0.6968 |
| **Asian subpopulation** | | | | |
| Alcohol use disorder | 0.89 (0.30-2.65) | 0.8359 | 1.33 (0.76-2.34) | 0.3166 |
| **Age (referent = 65-74)** |  |  |  |  |
| 76-100 | 0.66 (0.17-2.55) | 0.5435 | 1.38 (0.74-2.54) | 0.3086 |
| **Comorbidity** |  |  |  |  |
| Cerebrovascular disease | 0.94 (0.20-4.39) | 0.9366 | 1.63 (0.81-3.28) | 0.1674 |
| Depression | 3.24 (1.06-9.90) | 0.0394 | 2.09 (0.88-4.93) | 0.0939 |
| Diabetes | 0.77 (0.22-2.73) | 0.6893 | 1.60 (0.87-2.94) | 0.1293 |
| Fall | 0.67 (0.08-5.49) | 0.7063 | 0.61 (0.08-4.46) | 0.6236 |
| Heart disease | 1.78 (0.34-9.44) | 0.4989 | 0.55 (0.19-1.60) | 0.2722 |
| Hypertension | 1.51 (0.31-7.36) | 0.6092 | 0.60 (0.30-1.21) | 0.1546 |
| Liver disease | 1.70 (0.50-5.75) | 0.3908 | 0.73 (0.31-1.74) | 0.4827 |
| Chronic pulmonary disease | 0.92 (0.27-3.21) | 0.9005 | 1.24 (0.66-2.35) | 0.5079 |
| Renal disease | 0.50 (0.06-4.17) | 0.5189 | 0.61 (0.25-1.52) | 0.2907 |

**Table S4.** Demographics of alcohol abuse patient and matched controls.

|  | **Female** (N=42,164) | | | | **Male** (N=77,884) | | | |
| --- | --- | --- | --- | --- | --- | --- | --- | --- |
|  | Alcohol use disorder | | Matched Control | | Alcohol use disorder | | Matched Control | |
| **Age group** | | | | | | | | |
| 65-75 | 14,235 | 67.50% | 14,235 | 67.50% | 28,976 | 74.40% | 28,976 | 74.40% |
| 76-100 | 6,847 | 32.50% | 6,847 | 32.50% | 9,966 | 25.60% | 9,966 | 25.60% |
| **Race** |  |  |  |  |  |  |  |  |
| White | 16,209 | 76.90% | 16,209 | 76.90% | 28,234 | 72.50% | 28,234 | 72.50% |
| Hispanic | 1,384 | 6.60% | 1,384 | 6.60% | 3,787 | 9.70% | 3,787 | 9.70% |
| Black | 2,097 | 9.90% | 2,097 | 9.90% | 4,204 | 10.80% | 4,204 | 10.80% |
| Asian | 238 | 1.10% | 238 | 1.10% | 682 | 1.80% | 682 | 1.80% |
| Unknown | 1,154 | 5.50% | 1,154 | 5.50% | 2,035 | 5.20% | 2,035 | 5.20% |
| **Comorbidity** |  |  |  |  |  |  |  |  |
| Cerebrovascular disease | 4,272 | 20.30% | 4,272 | 20.30% | 7,455 | 19.10% | 7,455 | 19.10% |
| Depression | 7,752 | 36.80% | 7,752 | 36.80% | 6,446 | 16.60% | 6,446 | 16.60% |
| Diabetes | 4,795 | 22.70% | 4,795 | 22.70% | 11,887 | 30.50% | 11,887 | 30.50% |
| Fall | 4,022 | 19.10% | 4,022 | 19.10% | 3,738 | 9.60% | 3,738 | 9.60% |
| Heart disease | 3,435 | 16.30% | 3,435 | 16.30% | 8,229 | 21.10% | 8,229 | 21.10% |
| Hypertension | 16,265 | 77.20% | 16,265 | 77.20% | 30,481 | 78.30% | 30,481 | 78.30% |
| Liver disease | 2,297 | 10.90% | 2,297 | 10.90% | 3,833 | 9.80% | 3,833 | 9.80% |
| Chronic pulmonary disease | 8,393 | 39.80% | 8,393 | 39.80% | 12,933 | 33.20% | 12,933 | 33.20% |
| Renal disease | 3,025 | 14.30% | 3,025 | 14.30% | 5,785 | 14.90% | 5,785 | 14.90% |
| Traumatic brain injury | 358 | 1.70% | 358 | 1.70% | 314 | 0.80% | 314 | 0.80% |
